# Can large language models approximate human perceptions of disease severity? An evaluation using Global Burden of Disease 2010 disability weights

**DOI:** 10.64898/2026.05.02.26352261

**Authors:** Yeonju Ha, Hyewon Park, Yubin Lee, Siho Kim, Sangzin Ahn

**Affiliations:** Cardiovascular and Metabolic Diseases Medical Research Center, Inje University College of Medicine, Busan, Korea; Department of Pharmacology and Institute of Pharmacogenomics and Precision Medicine, Inje University College of Medicine, Busan, Korea

**Keywords:** Large Language Models, Global Burden of Disease, Disability-Adjusted Life Years, Disability Evaluation, Attitude to Health, Cross-Cultural Comparison, Post-Acute COVID-19 Syndrome

## Abstract

**Background:** Disability weights (DWs) quantify the severity of health loss and are essential for estimating disability-adjusted life years in the Global Burden of Disease (GBD) framework. Conventional DW estimation relies on resource-intensive population surveys that are difficult to update or adapt to emerging health states. Large language models (LLMs) may offer a scalable alternative by approximating human perceptions of disease severity through structured judgment tasks.

**Methods:** This exploratory study evaluated the alignment between LLM-derived and human-derived DW rankings using 222 health states from GBD 2010. All possible pairwise comparisons (24,531 pairs, each repeated three times) were conducted across four LLMs (GPT-5 mini, GPT-5, Claude Haiku 4.5, and Claude Sonnet 4.5). DWs were estimated via probit regression and evaluated using Spearman’s rank correlation and Steiger’s z test. The effects of prompt language (English vs. Korean), cultural role prompting, and medical specialist role prompting on alignment were examined. Additionally, the Binomial-Logit Indifference-Point (BLIP) estimator was proposed and validated through leave-one-out cross-validation for estimating DWs for health states without established values.

**Results:** All four LLMs showed high rank correlation with GBD 2010 DWs (Spearman’s ρ = 0.893 to 0.909), with no significant inter-model differences. Korean-language prompting significantly improved alignment with Korean DWs (ρ = 0.756 vs. 0.715, p = 0.011), and Korean cultural role prompting improved alignment with both GBD 2010 DWs (ρ = 0.922 vs. 0.909, p = 0.002) and Korean DWs (ρ = 0.738 vs. 0.715, p = 0.001). Medical specialist role prompting significantly reduced alignment with GBD 2010 DWs (ρ = 0.895 vs. 0.909, p = 0.001). BLIP demonstrated strong agreement with GBD 2010 DWs (Pearson’s r = 0.862, MAE = 0.066) and produced plausible estimates for Long COVID (mild: 0.020, moderate: 0.298, severe: 0.529).

**Conclusions:** LLMs can approximate human perceptions of disease severity with high rank-order consistency. Prompt language and role framing significantly influenced alignment, with culturally grounded lay prompting enhancing and specialist prompting reducing correspondence with population-based DWs. BLIP provides a practical framework for generating provisional DW estimates for emerging or underrepresented health states when conventional surveys are infeasible.

## 1. Background

Disability weights (DWs) are a core metric in epidemiological research for quantifying the severity of health loss associated with specific health states [1]. As a key component of disability-adjusted life years, DWs translate the burden of illness into a standardized, cross-condition severity scale. In the Global Burden of Disease (GBD) 2010 study, these weights were derived through large-scale household surveys conducted across five countries with approximately 14,000 respondents using pairwise comparison (PC), as well as through extensive web-based surveys across 167 countries with approximately 17,000 respondents that incorporated both PC and population health equivalence (PHE). These approaches were designed to capture generalized societal valuations of health state severity [2]. Importantly, DWs are intended to reflect social valuation by the general public rather than professional clinical judgment, a distinction that bears directly on how they should be estimated and interpreted.

However, these survey-based approaches are resource-intensive and difficult to update rapidly. The organizational and financial demands of large-scale valuation studies limit their responsiveness to evolving clinical and public health contexts, creating evidence gaps for emerging or changing conditions for which no established DWs exist. National adaptations, such as a Korean DW study [3], face similar challenges, requiring independent survey infrastructure to reflect local population values. The practical limitations of DW derivation thus represent a recognized challenge for burden of disease estimation, motivating the search for complementary approaches.

Meanwhile, large language models (LLMs) have shown promise across several medical tasks, including clinical note summarization [4], diagnostic support from radiology reports [5], and detection of depression and suicide risk from patient narratives [6]. Beyond these expert-oriented applications, LLMs may also have the potential to approximate human perceptions of disease severity through structured judgment tasks. Recent work [7] suggests that LLM-based generative agents can model healthcare decision-making and reproduce survey-like responses in silico, raising the possibility of estimating DWs for conditions lacking established values. This potential, however, raises important questions regarding validity and robustness. In particular, the alignment between LLM-derived and human-derived DWs requires evaluation. Furthermore, because LLM outputs may be shaped by prompt language and cultural framing [8], as well as by role prompting or in-context impersonation [9], it is necessary to evaluate the sensitivity of these value judgments to such conditions.

This study therefore addressed three objectives. First, we evaluate the alignment between LLM-derived DW rankings and GBD 2010 DW rankings across both OpenAI GPT and Anthropic Claude model families. Second, we examine how prompt conditions, including prompt language, cultural role prompting, and medical specialist role prompting, influence this alignment. Third, we propose and validate the Binomial–Logit Indifference-Point estimator (BLIP), a dedicated LLM-based framework for estimating DWs for conditions without established values. This was designed to overcome the temporal, resource, and infrastructure constraints of conventional population surveys. Taken together, these analyses address both the conceptual alignment between LLMs and human perception of disease severity and the practical utility of LLMs as scalable supplementary tools for DW estimation.

## 2. Methods

### 2.1 Study Design and Data Sources

This was an exploratory study evaluating the alignment between LLM-based and human-based perceptions of disease severity, using GBD 2010 as the primary reference standard. From the GBD 2010 appendix, 220 health states and their lay descriptions were obtained. Two anchoring states, “full health” and “being dead,” were added to define the boundary conditions of the disability weight scale, resulting in 222 health states. For analyses examining the influence of prompt language and cultural role prompting, DWs from a Korean population study were additionally incorporated, with the corresponding Korean lay descriptions adopted for the same 222 health states.

GBD 2010 DWs were estimated through a combination of PC and PHE, while the aforementioned Korean study demonstrated that PC alone yielded closely comparable estimates. The present study therefore adopted this simpler PC-only framework for deriving LLM-based DWs. Because the two approaches may still produce different value distributions, all primary analyses relied on rank-based measures, including Spearman’s rank correlation and rank differences, which capture the hierarchical structure of disease severity perception rather than depending on raw DW magnitudes.

### 2.2 Alignment between LLMs and Humans

#### 2.2.1 Base Prompt Design and Parameter Configuration

Considering that the GBD 2010 study was conducted among the general public, the prompts instructed the LLMs to respond from the perspective of a layperson without a medical background. For each PC, LLMs were instructed to read the lay descriptions of the two individuals’ health states and then (1) provide a brief rationale for the judgment in a sentence and (2) select the healthier one. This sequential response format was used to encourage more deliberate comparisons while preserving interpretability of the model’s judgments. This base prompt was developed in English with a fixed output format (Additional file 1). The definition of health used in the prompt was based on GBD 2010 and the World Health Organization (WHO) [10].

#### 2.2.2 Model-Based Pairwise Comparison and DW Derivation

To estimate LLM-based DWs, all possible pairs among the 222 health states, totaling 24,531 pairs, were constructed, and each comparison was repeated three times. A probit regression model was fitted to the binary choice results, in which the LLM selected the healthier of the two health states, following established discrete-choice methods used in health valuation research [11]. These coefficients were then anchored to a 0–1 scale, with “full health” defined as 0 and “being dead” defined as 1, consistent with the GBD 2010 framework. Based on this procedure, LLM-based DWs were estimated for four models: GPT-5 mini, GPT-5, Claude Haiku 4.5, and Claude Sonnet 4.5. Spearman’s ρ was used to evaluate the similarity between each model-derived DW ranking and the GBD 2010 DW ranking, as well as inter-model agreement. Subsequently, Steiger’s z test was applied to assess the statistical significance of differences between dependent correlation coefficients [12].

Within each disease group defined in the GBD 2010 appendix, LLM-based DW rankings were compared with GBD 2010 DW rankings for each model. A Wilcoxon signed-rank test was conducted to evaluate whether the rank differences were statistically significant. Finally, to examine model-specific patterns at the individual disease level, the rank differences between LLM-based and GBD 2010 DW rankings were calculated for each model. The top 10 diseases that were most overestimated and most underestimated by LLMs relative to humans were identified.

Subsequent analyses of prompt conditions and BLIP were conducted using GPT-5 mini alone, which was selected because it showed the highest observed alignment and favorable computational efficiency.

### 2.3 Analysis of Prompt Conditions Influencing Alignment and Significance Testing

To evaluate factors influencing the LLM’s perceptions of disease severity, the following factors were examined: prompt language, cultural role prompting, and medical specialist role prompting. Spearman’s ρ was calculated between DW rankings obtained under different prompt conditions and either GBD 2010 DW rankings or Korean DW rankings. Furthermore, Steiger’s z test was used to assess the statistical significance of differences between the correlations obtained under each prompt condition and those obtained under the base prompt, using the same reference ranking.

#### 2.3.1 Prompt Language

The base English prompt was translated into Korean (Additional file 2), and DWs were calculated following the same procedure. Spearman’s ρ was calculated for DW rankings obtained from the base and Korean prompts against both GBD 2010 and Korean DW rankings, and the resulting correlations were compared.

#### 2.3.2 Cultural Role Prompting

DWs were calculated under a condition in which a Korean identity was added to the base English prompt (Additional file 3). Spearman’s ρ was calculated for DWs obtained from both the base prompt and the Korean role condition against GBD 2010 and Korean DW rankings, respectively. Steiger’s z test was then performed to evaluate differences in alignment according to role prompting, using GBD 2010 DWs and Korean DWs as the respective reference standards.

#### 2.3.3 Medical Specialist Role Prompting

DWs were calculated under a condition where a medical specialist role was assigned to the base English prompt (Additional file 4). Spearman’s ρ was calculated for DW rankings obtained under both conditions against GBD 2010 DW rankings. Differences in alignment were evaluated using Steiger’s z test.

### 2.4. BLIP

In addition to the alignment analyses, this study evaluated the practical utility of LLM-based pairwise judgments for estimating DWs for health states without established values. For this purpose, the Binomial–Logit Indifference-Point estimator (BLIP) was developed, validated, and applied.

#### 2.4.1 BLIP Framework

The BLIP method estimates DWs for health states without established values by adapting the classical psychophysical method of constant stimuli, using the concept of the Point of Subjective Equality (PSE) as its central metric. In this context, PSE is the point at which two stimuli are perceived as equivalent, yielding equal choice probabilities [13].

Repeated PCs were conducted using an LLM between a target disease (U), whose DW was to be estimated, and multiple anchor diseases (k) with known DWs. For each pair, the LLM was repeatedly asked to judge which condition was healthier, and the resulting win-loss counts were recorded.

These data were modeled using a binomial generalized linear model with a logit link function. Let p_k_ denote the model-estimated probability that U is selected as healthier than anchor disease k, and let DW_k_ denote the known DW of anchor disease k. The model is formulated as follows:

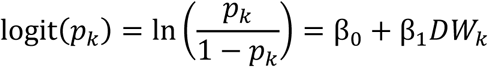

As the severity of the anchor disease (DW_k_) increases, U becomes more likely to be judged as the healthier condition, so β_1_ > 0 is expected.

The estimated DW_U_ was then obtained as the PSE of the fitted model, which represents the value of DW_k_ at which the predicted selection probability (p_k_) equals 0.5. At this point of indifference, where logit(p_k_) = 0, the estimated weight is as follows:

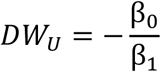

#### 2.4.2 Validation of BLIP

To evaluate the reliability of BLIP, leave-one-out validation was performed across the 220 GBD 2010 health states. In each iteration, one health state was treated as the target, and its DW was estimated using the remaining 221 states as anchors, comprising the other 219 GBD 2010 health states together with the two fixed anchoring states of full health and being dead. The estimated values were then compared with the corresponding GBD 2010 DWs. Performance was evaluated using mean absolute error (MAE), mean bias, and Pearson’s correlation coefficient (r).

#### 2.4.3 Application of BLIP

To demonstrate the applicability of BLIP to newly emerging conditions, DWs for Long COVID, which is not included in the GBD 2010, were estimated. Lay descriptions from the GBD 2010 study were provided to GPT as stylistic references, together with a review article on COVID-19-associated neurological and psychological manifestations [14]. GPT was then prompted to generate lay descriptions for mild, moderate, and severe Long COVID. The resulting descriptions are presented in Additional file 5.

## 3. Results

### 3.1 Overall Alignment between LLM-based DWs and GBD 2010 DWs

The LLM-based DWs derived using the base prompt showed high correlation with GBD 2010 DWs across all models. Spearman’s ρ values were 0.909 for GPT-5 mini, 0.902 for GPT-5, 0.898 for Claude Haiku 4.5, and 0.893 for Claude Sonnet 4.5. Steiger’s test revealed no statistically significant differences in any pairwise model comparison, indicating comparable alignment across models (Additional file 6). These findings suggest that alignment with GBD 2010 DWs was similar across the evaluated LLMs. Furthermore, a high degree of agreement was also observed among DWs derived from different LLMs (Additional file 7). A comparison of rank distributions between GBD 2010 DWs and LLM-based DWs by disease group is presented in Figure 1.

**Figure 1.**
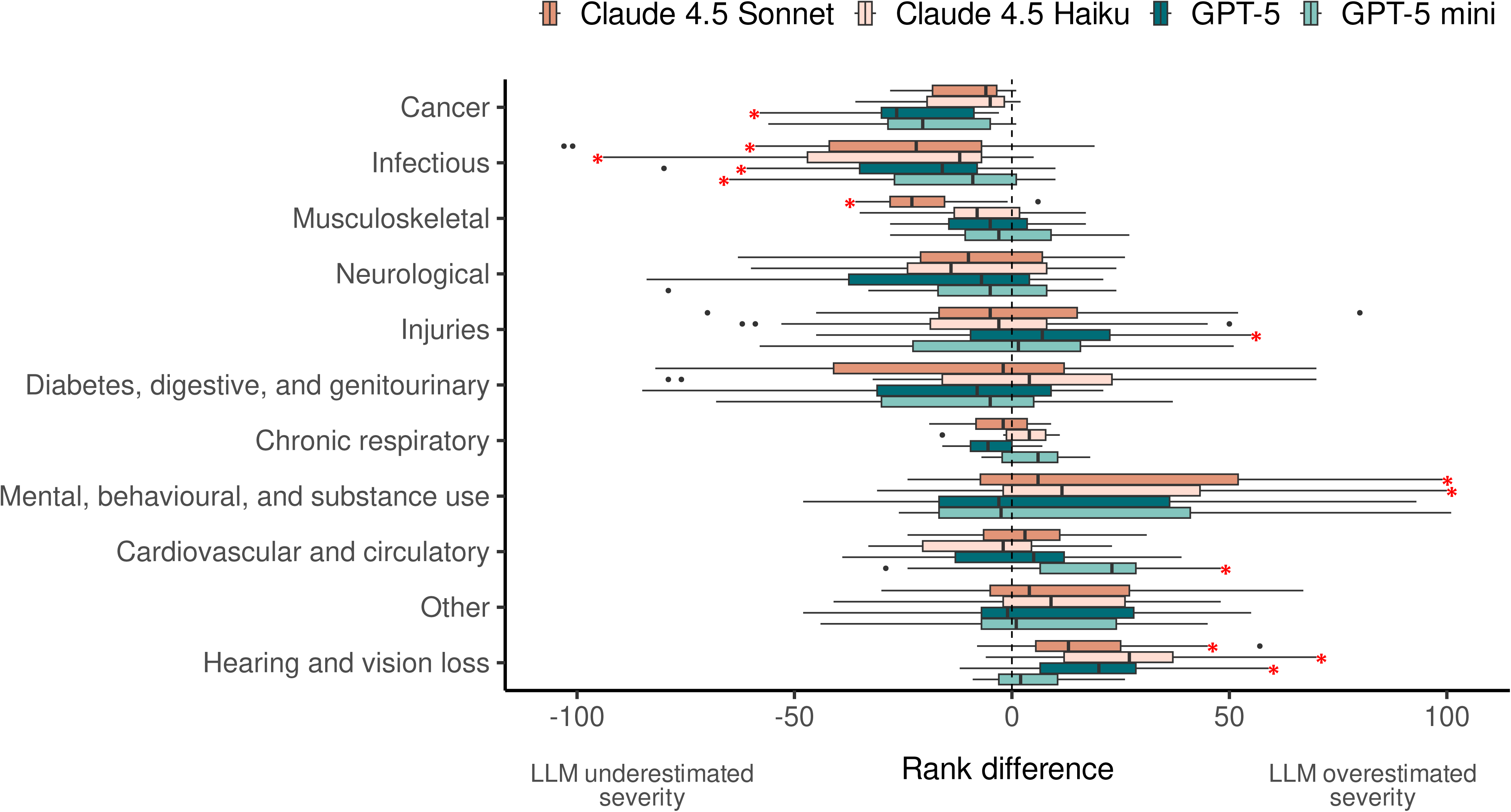
Rank differences between GBD 2010 and LLM-based disability weights by disease group. Box plots showing the distribution of rank differences (LLM-based DW rank minus GBD 2010 DW rank) for each disease group across four LLMs (GPT-5 mini, GPT-5, Claude Haiku 4.5, and Claude Sonnet 4.5). Positive values indicate that the LLM overestimated disease severity relative to GBD 2010, and negative values indicate underestimation. Asterisks denote statistically significant rank differences within a disease group (Wilcoxon signed-rank test, p < 0.05).

At the individual disease level, LLMs consistently underestimated the severity of Crohn’s disease or ulcerative colitis, moderate and severe diarrhea, and epididymo-orchitis relative to GBD 2010. In contrast, LLMs consistently overestimated the severity of moderate and severe fetal alcohol syndrome and moderate, severe, and profound intellectual disability. Detailed results for the top 10 most underestimated and overestimated diseases across all models are shown in Tables 1 and 2.

**Table 1.**
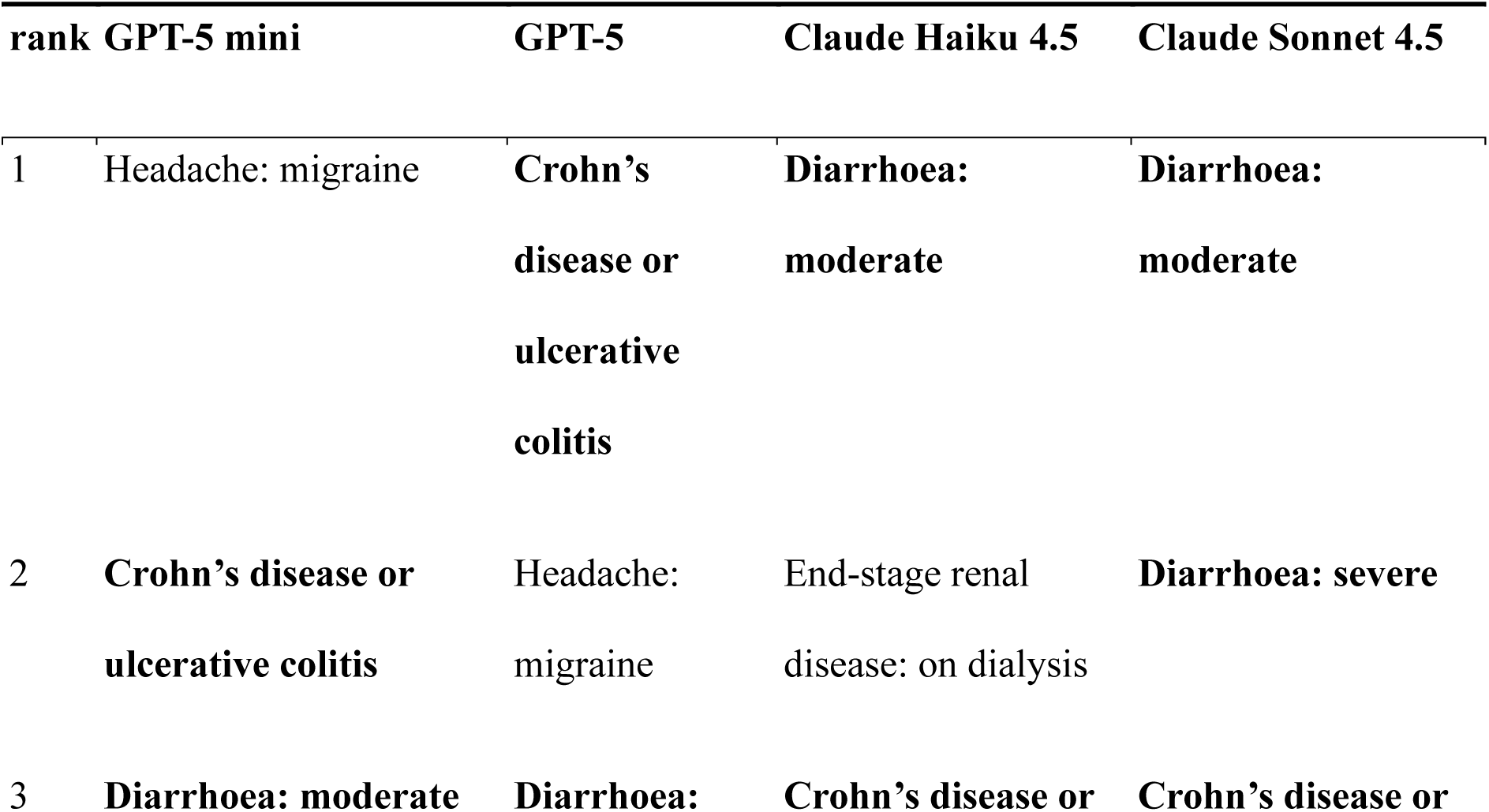
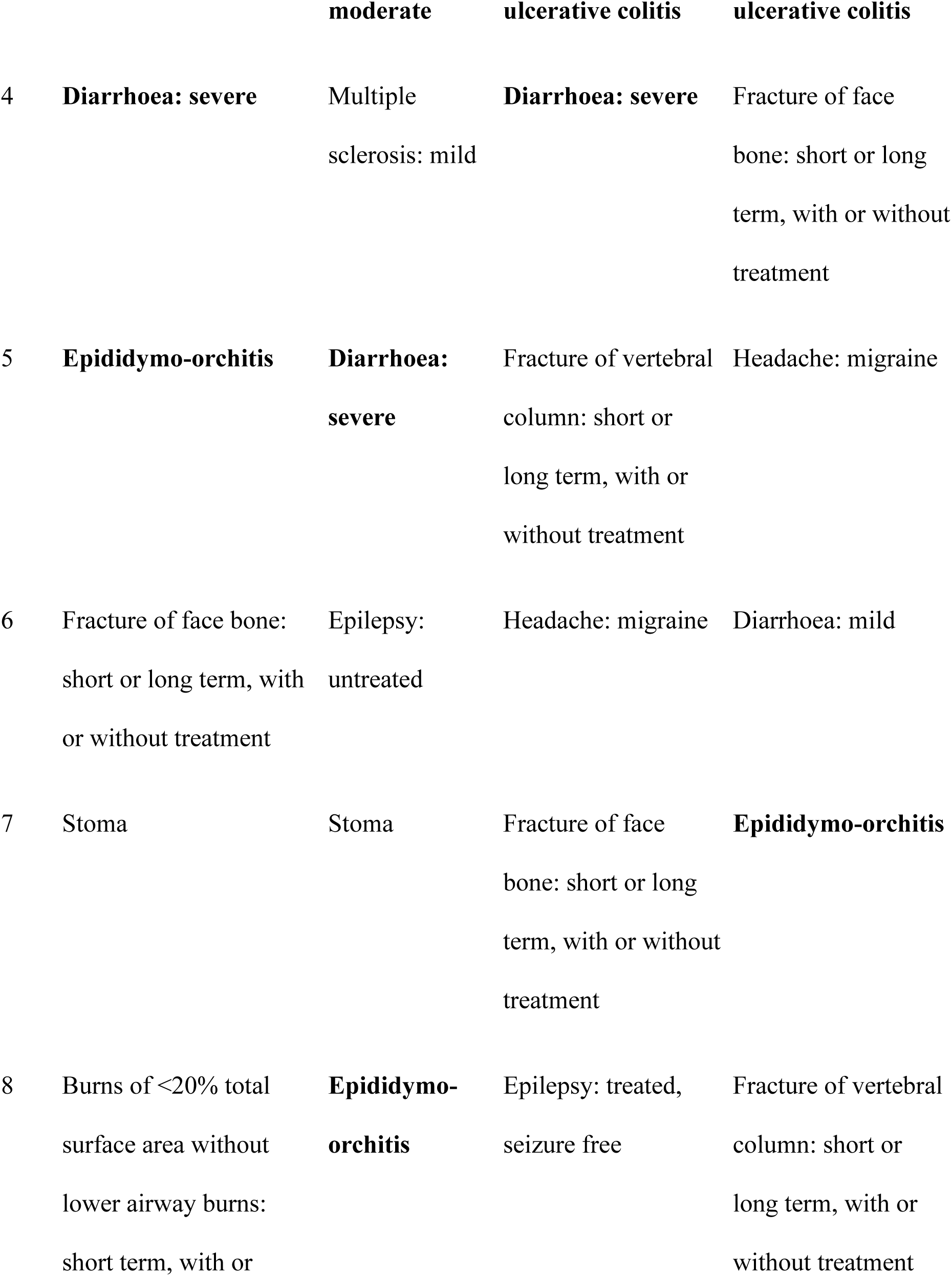
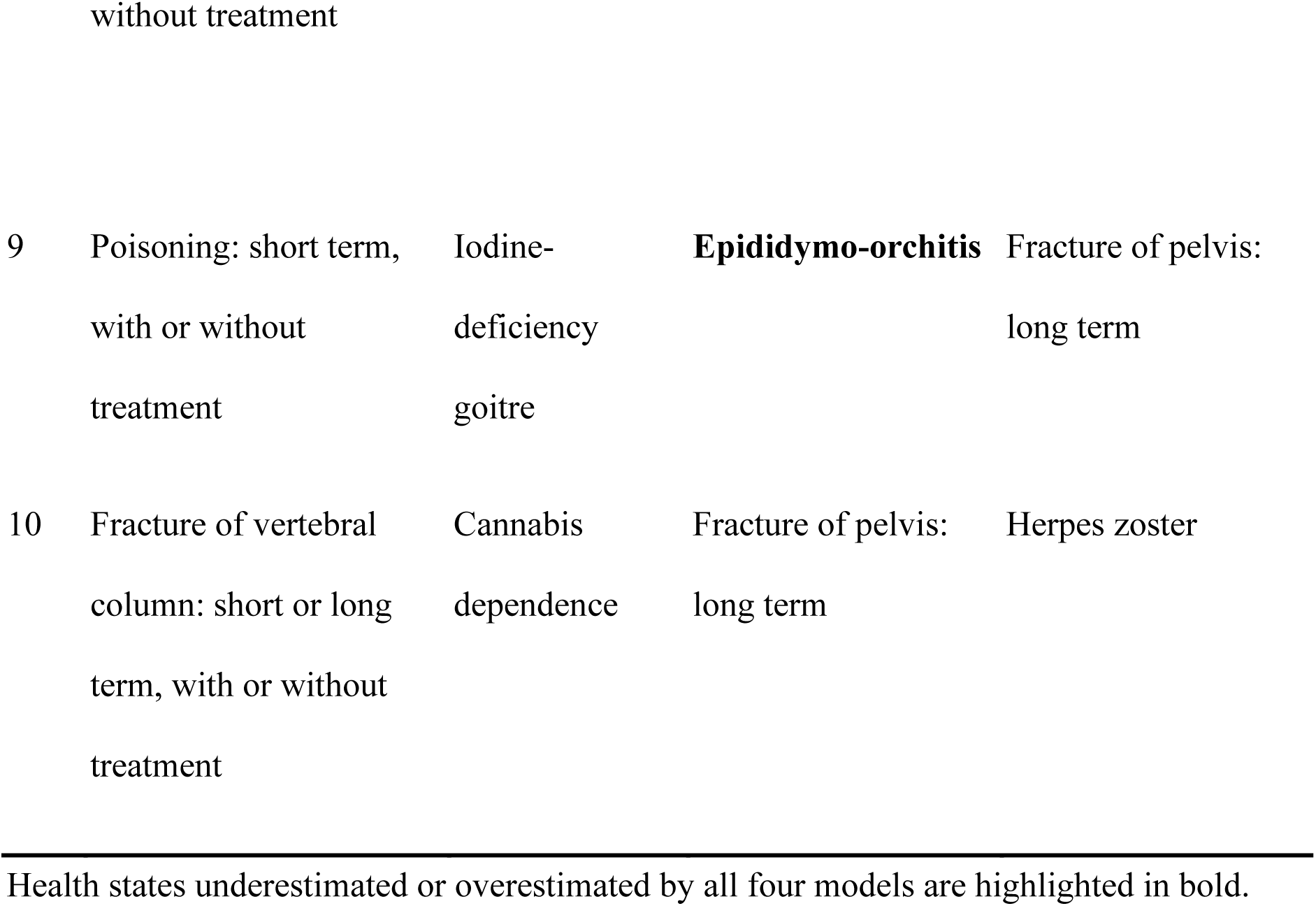
Top 10 LLM-Underestimated Health States Identified by DW Ranking Differences.

**Table 2.** Top 10 LLM-Overestimated Health States Identified by DW Ranking Differences.

| rank | GPT-5 mini | GPT-5 | Claude Haiku 4.5 | Claude Sonnet 4.5 |
| --- | --- | --- | --- | --- |
| 1 | **Intellectual disability: moderate** | **Intellectual disability: severe** | **Intellectual disability: moderate** | **Intellectual disability: moderate** |
| 2 | **Intellectual**<br>**disability: severe** | **Intellectual**<br>**disability:**<br>**profound** | **Intellectual**<br>**disability: profound** | **Intellectual**<br>**disability:**<br>**severe** |
| 3 | **Intellectual**<br>**disability: profound** | **Intellectual**<br>**disability:**<br>**moderate** | **Intellectual**<br>**disability: severe** | **Intellectual**<br>**disability:**<br>**profound** |
| 4 | **Fetal alcohol**<br>**syndrome:**<br>**moderate** | **Fetal alcohol**<br>**syndrome:**<br>**severe** | Hearing loss:<br>complete | Spinal cord<br>lesion below<br>neck: treated |
| 5 | **Fetal alcohol**<br>**syndrome: severe** | Hearing loss:<br>profound | Infertility: primary | Intellectual<br>disability: mild |
| 6 | Autism | **Fetal alcohol**<br>**syndrome:**<br>**moderate** | Hearing loss:<br>profound | Infertility:<br>primary |
| 7 | Spinal cord lesion<br>below neck: treated | Severe wasting | **Fetal alcohol**<br>**syndrome:**<br>**moderate** | **Fetal alcohol**<br>**syndrome:**<br>**moderate** |
| 8 | Asperger's syndrome | Spinal cord<br>lesion below | Infertility: secondary | Motor plus<br>cognitive |
|  |  | neck: treated |  | impairments:<br>mild |
| 9 | Stroke: long-term<br>consequences,<br>moderate | Intellectual<br>disability: mild | **Fetal alcohol**<br>**syndrome: severe** | **Fetal alcohol**<br>**syndrome:**<br>**severe** |
| 10 | Dislocation of knee:<br>long term, with or<br>without treatment | Autism | Amputation of<br>finger(s), excluding<br>thumb: long term,<br>with treatment | Hearing loss:<br>profound |

Health states underestimated or overestimated by all four models are highlighted in bold.

### 3.2 Factors Influencing Alignment

Based on the preceding model comparison, GPT-5 mini was selected for subsequent analyses because it showed the highest observed alignment together with favorable cost efficiency. The results of the analyses of factors influencing alignment are presented in Table 3.

**Table 3.**
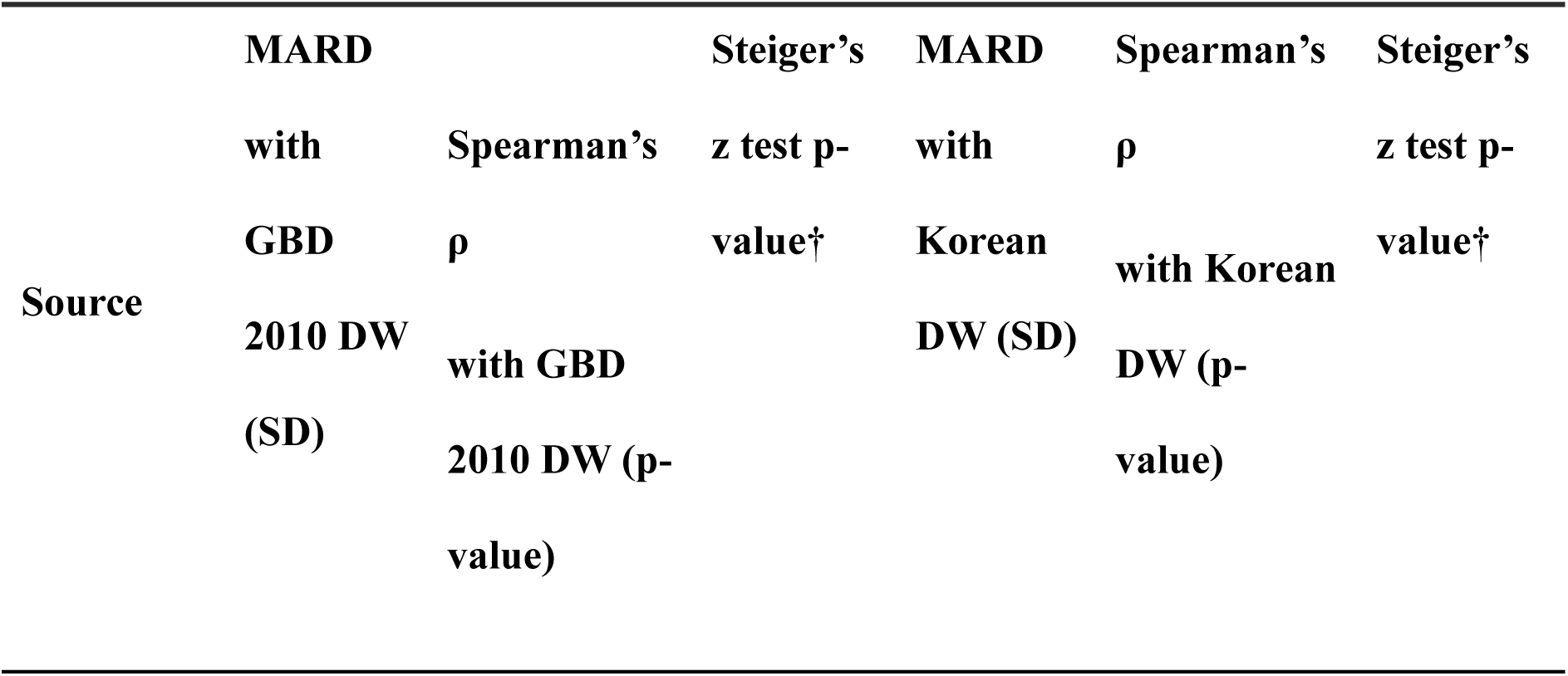
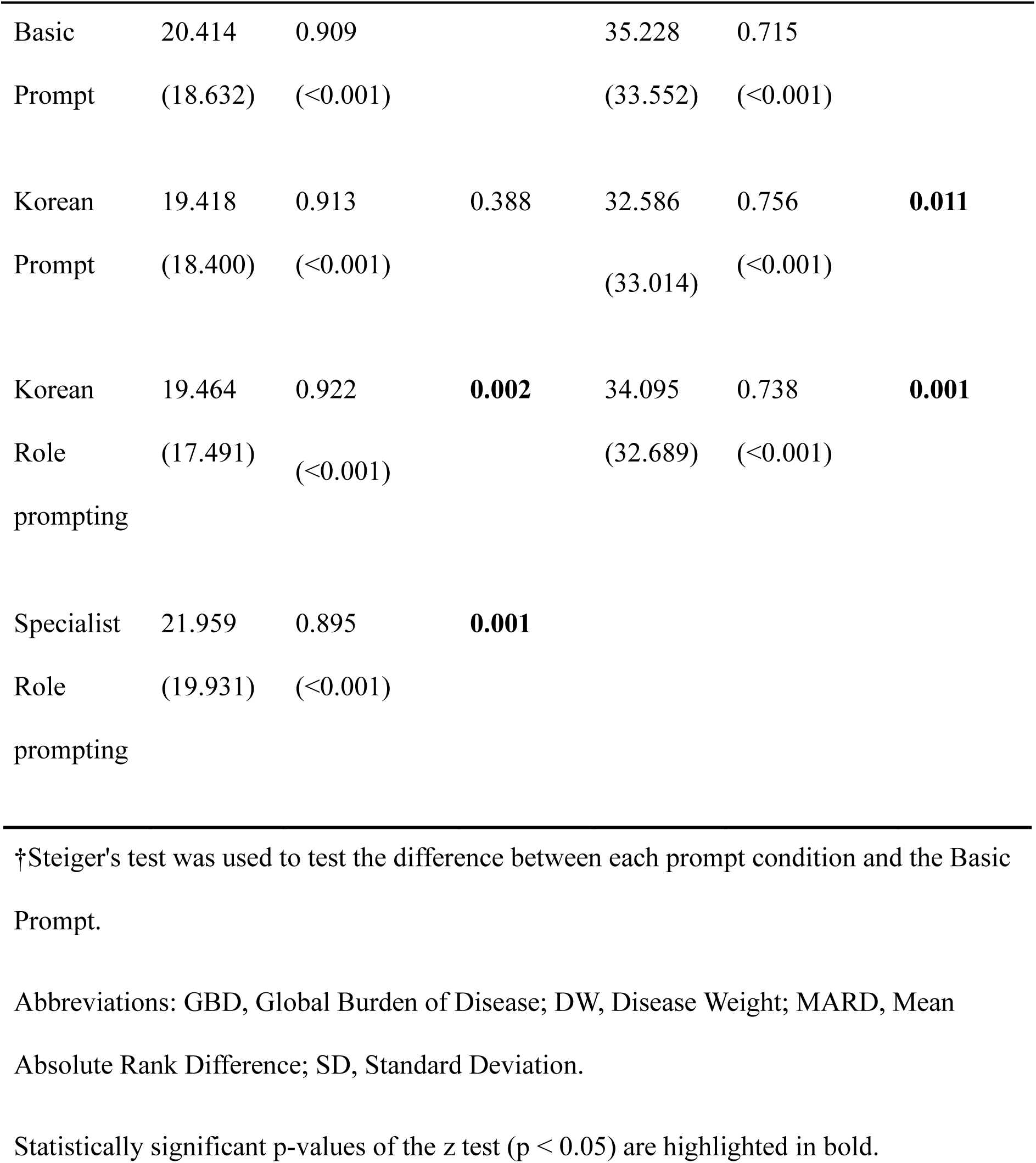
Comparison of Spearman’s ρ according to Prompt Language.

#### 3.2.1 Prompt Language

When compared with GBD 2010 DWs, no significant difference in Spearman’s ρ was observed between the base and Korean prompts (0.909 vs. 0.913, Steiger’s z test, p = 0.388). However, when Korean DWs were used as the reference, the Korean prompt yielded a significantly higher Spearman’s ρ than the base prompt (0.756 vs. 0.715, p = 0.011).

#### 3.2.2 Cultural Role Prompting

DWs derived from the Korean layperson role prompt exhibited significantly higher Spearman’s ρ than those derived from the base prompt against both GBD 2010 DWs (0.922 vs. 0.909, p = 0.002) and Korean DWs (0.738 vs. 0.715, p = 0.001).

#### 3.2.3 Medical Specialist Role Prompting

Conversely, DWs derived from a prompt assigning a medical specialist role showed a significantly lower Spearman’s ρ with GBD 2010 DWs than those derived from the base prompt (0.895 vs. 0.909, p = 0.001).

### 3.3 BLIP

#### 3.3.1 Overall Performance of BLIP

Across 220 health states, BLIP-estimated DWs showed strong agreement with GBD 2010 DWs (Figure 2): Pearson’s r = 0.862, MAE = 0.066, and bias = 0.023.

**Figure 2.**
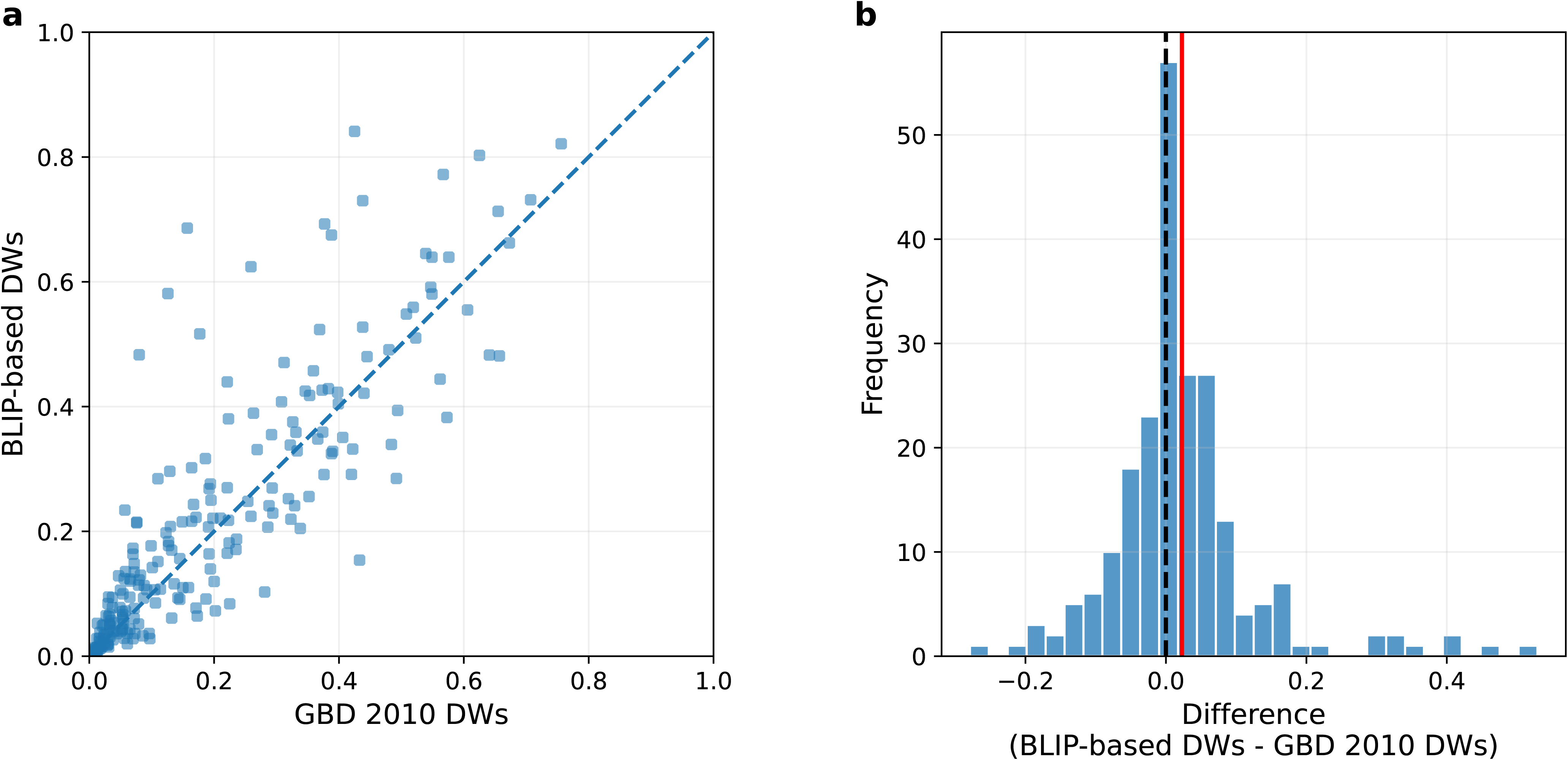
BLIP-based estimation of disability weights. (a) Scatter plot of GBD 2010 DWs (x-axis) versus BLIP-estimated DWs (y-axis) across 220 health states. The dashed diagonal line denotes perfect agreement. Pearson’s r, mean bias, and MAE are shown. (b) Histogram of the differences between BLIP-estimated and GBD 2010 DWs. The black dashed line indicates zero difference; the red solid line indicates the mean error (+0.023).

#### 3.3.2 Long COVID DW Prediction

BLIP-estimated DWs for long COVID were 0.020 (mild), 0.298 (moderate), and 0.529 (severe).

## 4. Discussion

### 4.1 Summary and Interpretation of the Results

This study showed that LLM-derived DW rankings were highly aligned with GBD 2010 DWs across all four evaluated models. Despite differences in model families, all models produced similarly strong correlations with the GBD 2010 reference, and no statistically significant between-model differences were observed. These findings suggest that LLMs can approximate human perceptions of disease severity when judgments are based on standardized lay descriptions of health states. Although the GBD 2010 DWs were derived using a combination of PC and PHE, whereas our LLM-based estimates were based on PC alone, the strong rank-based alignment indicates that LLMs were able to reproduce the relative ordering of health-state severity with considerable consistency.

High agreement was also observed among DW rankings generated by different LLMs. This suggests that, despite architectural and training differences, these models may encode broadly similar patterns of disease-severity valuation. Such convergence supports the possibility that LLMs capture aspects of shared social knowledge embedded in language, rather than producing highly idiosyncratic judgments specific to a particular model family. At the same time, the present findings do not imply that LLMs fully replicate human judgment in all respects. Rather, they suggest that collective societal perceptions of disease severity are substantially encoded in language and recoverable through LLMs.

Although the overall distributions of LLM-based and GBD 2010 DW rankings were largely comparable, several discrepancies were shared across models. LLMs tended to underestimate the severity of conditions such as diarrhea and epididymo-orchitis, while overestimating the severity of fetal alcohol syndrome and intellectual disability. One possible interpretation is that disease-severity judgments reflect both a language-recoverable component and an experience-dependent component. LLMs may more readily capture conditions whose severity is strongly expressed in language through chronicity, lifelong developmental consequences, or strong negative social valence, while underrepresenting conditions whose burden depends more heavily on lived bodily discomfort, embarrassment, or disruptions to everyday functioning. This interpretation remains heuristic rather than directly demonstrated, but it is consistent with recent evidence [15] that language-only LLMs align more closely with humans on non-sensorimotor than on sensorimotor dimensions.

Prior studies report conflicting evidence on prompt language effects: Alkhamissi et al. [16] found that native-language prompts shifted outputs toward corresponding cultural values, whereas Cao et al. [17] observed inconsistent or reversed effects depending on the cultural dimension measured. Our results partially support both findings. Korean prompts did not alter alignment with the multinational GBD 2010 reference, but significantly improved alignment with Korean-specific DWs, suggesting that prompt language selectively enhances culturally proximate value judgments rather than uniformly shifting all outputs. Nevertheless, given the conflicting prior findings, further investigation is needed as the effect of language on alignment may vary depending on the task or the language.

Meanwhile, assigning a Korean identity was associated with improved alignment with both GBD 2010 DWs and Korean DWs. Given that GBD is based on multinational surveys to minimize country-specific value biases and ensure generalizability, this improvement suggests that assigning a Korean identity may partially attenuate U.S.-centric value biases amplified in English prompts similar to the prior research [18]. Consequently, such cultural framing may induce a more universally applicable structure of value judgment. Furthermore, improved alignment with Korean DWs under a Korean identity supports that a cultural role can align model judgments more closely with the particular cultural context. This finding aligns with Tao et al. [19], which demonstrated that cultural role assignments improved alignment with region-specific survey responses across multiple countries, suggesting that the effect observed here is not unique to Korean contexts.

Under the medical specialist identity, the alignment with GBD 2010 DWs significantly decreased. This phenomenon may be interpreted as a shift in judgment criteria toward factors such as disease prognosis, pathophysiological mechanisms, or epidemiological indicators. Such a shift can diverge from evaluations grounded in social dimensions, including perceived limitations in daily life and impairments in social functioning, experienced subjectively by patients and the general population. Accordingly, assigning a specialist identity may have produced results that are incompatible with the nature of DWs as measures of social value.

BLIP showed reasonable agreement with GBD 2010 DWs across 220 health states, suggesting that this method can generate estimates broadly consistent with established DWs. For long COVID, BLIP-estimated DWs were 0.020, 0.298, and 0.529 for mild, moderate, and severe states, respectively, and their monotonic increase supports ordinal face validity. Although long COVID DWs have not been directly estimated in population surveys, a study mapping long COVID symptom clusters to existing GBD health states [20] reported DWs of 0.02 to 0.41 for respiratory symptoms, 0.07 to 0.38 for cognitive problems, and 0.22 for persistent fatigue with pain. Because our long COVID descriptions represent composite states across symptom domains, the BLIP estimates fall within or near these ranges, providing preliminary external support for their plausibility. Although long COVID is presented here as an illustrative example, this approach could also be extended to health states without existing DWs, including individualized health-state descriptions generated on demand. More broadly, BLIP can convert any target health state description into DW estimates calibrated to a given reference scale, enabling alignment with country-specific or international DW systems as needed.

### 4.2 Implications

According to prior study [21], differences between clinicians’ and laypersons’ perceptions of disease can affect patient satisfaction and treatment adherence. In this study, LLMs showed statistically meaningful alignment with lay perceptions of health states. If validated in clinical settings, LLM-generated lay perspectives could serve as a preparatory aid for patient communication by helping clinicians anticipate gaps between clinical and public perceptions of disease severity. However, infectious diseases tended to be underestimated, so caution is warranted in this domain.

This study also provides a basis for future work on LLM-human perceptual alignment by identifying prompt-related factors that may influence it. Because alignment was comparable across models, future studies may reasonably prioritize cost-effectiveness when selecting models.

From a health policy perspective, BLIP may offer a practical supplementary approach for estimating DWs in settings where traditional valuation studies are difficult to conduct. Conventional DW estimation requires extensive survey infrastructure, time, and financial resources. In contrast, the total API cost per model ranged from $8.58 to $78.27 for three repeated runs across all 24,531 pairs, whereas the multi-country household surveys underlying GBD 2010 required thousands of respondents and substantial logistical support. Although these approaches are not directly comparable in scope or evidentiary status, their resource demands differ by several orders of magnitude. In this context, BLIP may be particularly useful for emerging conditions, rare diseases, or other situations in which timely provisional estimates are needed before formal survey-based DWs become available.

### 4.3 Limitations

This study has several limitations. First, although systematic overestimation and underestimation were observed for specific disease groups and individual health states, the underlying reasons could not be directly established. The model-generated rationales were often limited to symptom enumeration or generic paraphrases of health-state descriptions, and therefore did not reliably reveal the basis of the model’s judgments. As a result, qualitative interpretation of these rationales was limited. In addition, because the GBD study does not report the reasoning underlying respondents’ choices, direct comparison between human and LLM reasoning was not possible. Quantitative text-based approaches, including part-of-speech analysis and other text-mining methods, were also explored, but they did not provide sufficiently informative signals for identifying the mechanisms underlying the observed discrepancies.

Second, the analysis of prompt-related factors was limited in scope. Only English and Korean were examined, and role prompting was restricted to a generic layperson, a Korean layperson, and a medical specialist. Accordingly, the findings should not be generalized to other languages, cultural identities, or role configurations without further study. Additional work is needed to determine whether the observed prompt effects extend to broader linguistic and sociocultural settings.

Third, the alignment analyses used GBD 2010 as the reference standard because it remains the most recent GBD study with publicly available detailed lay descriptions of health states. However, disease perceptions may shift over time, and contemporary LLMs may reflect present-day language use and value patterns rather than those prevailing at the time of GBD 2010. Therefore, some discrepancies between LLM-based DWs and GBD 2010 DWs may reflect temporal differences in social valuation rather than model error alone.

### 4.4 Conclusions

This study demonstrates that LLMs can achieve a high degree of alignment with human-derived perceptions of disease severity, as reflected in strong rank correlations between LLM-based DWs and GBD 2010 DWs. This alignment was observed across multiple model families, but was also meaningfully influenced by prompt language and role framing. In particular, culturally grounded lay prompting improved alignment, whereas specialist prompting reduced it, indicating that the social perspective encoded in the prompt is important for approximating DW judgments.

In addition, BLIP showed that LLM-based pairwise judgments can be extended beyond alignment analysis to support provisional DW estimation for health states without established values. Although such approaches cannot replace population-based valuation studies, they may serve as useful supplementary tools when formal surveys are infeasible or unavailable. Overall, these findings support the potential of LLMs as scalable, flexible, and context-sensitive aids for research on disease valuation, while also underscoring the need for continued validation and careful prompt design.

## Data Availability

All data analyzed in this study are publicly available through the Global Burden of Disease (GBD) 2010 study and the Korean Disability Weight study. The specific prompts and supplementary health state descriptions used to generate the LLM-based data are contained within the manuscript and its additional files.

https://doi.org/10.1016/S0140-6736(12)61680-8

https://doi.org/10.1371/journal.pone.0162478

## List of abbreviations

(DWs): Disability weights
(GBD): Global Burden of Disease
(PC): pairwise comparison
(LLMs): Large Language Model
(PHE): population health equivalence Binomial-Logit
(BLIP): Indifference-Point estimator
(WHO): World Health Organization
(PSE): Point of Subjective Equality
(MAE): Mean Absolute Error
(MARD): Mean Absolute Rank Difference
(SD): Standard Deviation

## Declarations

Ethics approval and consent to participate

Not applicable

Consent for publication Not applicable

Availability of data and materials

The original data are available from the GBD 2010 study and the Korean DW study. All experiments are reproducible using the GBD 2010 study, Korean DW study, the prompts provided in Additional files 1–4 and the descriptions presented in Additional file 5.

## Competing interests

The authors declare no competing interests.

## Funding

This research was supported by the National Research Foundation of Korea (NRF), funded by the Korean government (MSIT) (RS-2025-02214129)

## Author’s contributions

Y.H., H.P., Y.L., and S.K. were responsible for data retrieval, curation, analysis, and the interpretation of the results. S.A. provided overall supervision of the study, including guidance on study design and analytical approach. All authors contributed to drafting and revising the manuscript, critically reviewed its content, and approved the final version for publication.

## Acknowledgements

Not applicable

## Additional files

### Additional file 1

docx

Prompt Design, the Base Prompt, and Experimental Conditions

In this study, a prompt was developed based on the methodology used in the GBD 2010 study to evaluate the alignment of LLMs with human perceptions of disease severity.

The base prompt was written in English and designed to assess disease severity from the perspective of a layperson without a medical background. The model was presented with descriptions of two health states (provided as [1] and {L2} in the prompt) and was instructed to provide a one-sentence rationale for which state was healthier, followed by a final selection. The temperature parameter was fixed at 1 across all experiments, corresponding to the default setting.

### Additional file 2

docx

Korean Prompt

To examine the effect of prompt language on the alignment, the base prompt was translated into Korean. Semantic distortion was minimized during the translation process, while the output-format instructions were kept identical to those of the base prompt.

### Additional file 3

docx

Role Prompting: Prompt for Assigning a Cultural Identity

To examine whether culturally specific values are reflected, a phrase assigning a Korean identity was added to the Role definition section of the system prompt. In the modified prompt, underlined portions indicate changes to the base prompt, while all other elements were kept identical.

### Additional file 4

docx

Role Prompting: Prompt Assigning a Specialist Role

To examine whether assigning an specialist role alters value judgments relative to those of the general public, the role was changed from a layperson to a medical specialist. In the modified prompt, underlined portions indicate changes to the base prompt, while all other elements were kept identical.

### Additional file 5

docx

Descriptions of Long COVID

The descriptions of mild, moderate, and severe long COVID were defined as follows.

### Additional file 6

docx

Steiger’s z test for Pairwise Correlation Comparisons Across LLMs

Steiger’s z test was conducted to evaluate whether Spearman’s ρ between LLM-based DWs and GBD 2010 DWs differed significantly across model pairs.

### Additional file 7

docx

Inter-Model Rank Correlation of DW Rankings

To assess the consistency of DW rankings across the four different models, pairwise Spearman’s ρ were calculated. All inter-model correlations were high, with coefficients greater than 0.945.

## Notes

### Competing Interest Statement

The authors have declared no competing interest.

### Author Declarations

The study analyzed publicly available human-derived data from the Global Burden of Disease (GBD) 2010 study. The disability weights and health state descriptions were obtained from the published appendix of the following study: Primary Source: Salomon JA, Vos T, Hogan DR, Gagnon M, Naghavi M, Mokdad A, et al. Common values in assessing health outcomes from disease and injury: disability weights measurement study for the Global Burden of Disease Study 2010. The Lancet. 2012;380(9859):2129-43. DOI/Link: https://doi.org/10.1016/S0140-6736(12)61680-8 Additionally, the study used comparative data from the Korean Disability Weight study: Secondary Source: Ock M, Ahn J, Yoon S-J, Jo M-W. Estimation of disability weights in the general population of South Korea using a paired comparison. PLoS One. 2016;11(9):e0162478. DOI/Link: https://doi.org/10.1371/journal.pone.0162478

