## Additional file 1 for "Can large language models approximate human perceptions of disease severity? An evaluation using Global Burden of Disease 2010 disability weights"

  prompt1 = f"""

\# Role

Judge and answer as an ordinary person with no medical background.

(Examples of medical background knowledge: knowledge of potential treatments, long-term outlook (prognosis), medical causes of diseases (pathophysiological mechanisms), medical statistics (epidemiology), etc.)

Carefully review the provided scenario and evaluation criteria, and judge and answer using only common sense and knowledge at the layperson level.

\# Scenario

- The first person: {L1}

- The second person: {L2}

\# Evaluation Criteria

- "Healthier" generally means having fewer overall limitations on what one can do in life, or being in a better state of well-being physically, mentally, and socially.

- Do not focus only on the scope of the problem (localized vs. systemic); focus on its overall impact on life.

- There is no single correct answer; we want your opinion.

- You must follow the specified answer format.

- The reasoning must be **one sentence**, and **simple repetition or paraphrasing of the scenario is prohibited**.

- Do not include unnecessary sentences; write only the reasoning and the answer.

\# Answer Format

Reasoning_basis:

Answer: 1. The first person or 2. The second person

"""

  prompt2 = f"""

\# Question

Between the first person ({L1}) and the second person ({L2}), who do you think is overall healthier?

"""
