## Additional file 2 for "Can large language models approximate human perceptions of disease severity? An evaluation using Global Burden of Disease 2010 disability weights"

  prompt1 = f"""

\# 역할

의학적 배경지식이 없는 일반인처럼 판단하고 답변하세요.

(의학적 배경 지식의 예: 잠재적 치료법, 장기적 전망(예후), 질환의 의학적 원인(병태생리학적 기전), 의학 통계(역학) 등에 대한 지식이 없음)

제공된 상황과 평가기준을 꼼꼼하게 확인하고, 오직 일반인 수준에서의 상식과 지식만으로 판단하고 답변하세요.

\# 상황

- 첫 번째 사람: {L1}

- 두 번째 사람: {L2}

\# 평가기준

- "더 건강하다"는 일반적으로 삶에서 할 수 있는 일에 대한 전반적인 제약이 더 적거나, 신체적·정신적·사회적으로 더 나은 웰빙 상태에 있음을 의미합니다.

- 문제의 범위(국소적vs전신적)에만 집중하지 말고, 삶의 전반적 영향에 초점을 두세요.

- 정답은 없으며, 당신의 의견을 듣고자 합니다.

- 반드시 지정된 답변 형식을 따르세요.

- 판단 근거는 **한 문장**으로 작성하며, **단순 반복이나 상황에서 표현만 바꾼 설명은 금지**합니다.

- 불필요한 문장은 포함하지 말고, 판단 근거와 답변만 작성하세요.

\# 답변 형식

판단 근거:

답변: 1. 첫 번째 사람 or 2. 두 번째 사람

  """

  prompt2 = f"""

\# 질문

첫 번째 사람({L1})과 두 번째 사람({L2}) 중에서 전반적으로 누가 더 건강하다고 생각하나요?

  """
