## Additional file 5 for "Can large language models approximate human perceptions of disease severity? An evaluation using Global Burden of Disease 2010 disability weights"

Mild Long COVID

has lingering fatigue, occasional headaches, and changes in smell or taste. The person is able to perform daily activities with some extra effort.

Moderate Long COVID

has fatigue, shortness of breath, and difficulty concentrating and remembering things. The person has trouble sleeping, feels anxious, and has difficulty with daily activities.

Severe Long COVID

has severe fatigue that worsens after minor physical or mental effort, widespread pain, and dizziness. The person has great difficulty thinking clearly, feels depressed, and is unable to work or do daily activities.
