## Additional file 7 for "Can large language models approximate human perceptions of disease severity? An evaluation using Global Burden of Disease 2010 disability weights"

| Spearman’s ρ | **GPT-5 mini** | **GPT-5** | **Claude Haiku 4.5** | **Claude Sonnet 4.5** |
| --- | --- | --- | --- | --- |
| GPT-5 mini | - | 0.971 | 0.946 | 0.954 |
| GPT-5 | - | - | 0.945 | 0.960 |
| Claude Haiku 4.5 | - | - | - | 0.968 |
| Claude Sonnet 4.5 | - | - | - | - |
